# Continuous tracking of aortic aneurysm diameter with peripheral pulse waves: a computational framework combining sequential Markov chain Monte Carlo with Kalman filtering

**DOI:** 10.64898/2026.02.09.26345911

**Authors:** Kiran Bhattacharyya

## Abstract

**Objective:** Abdominal aortic aneurysms (AAA) affect more than 1% of adults over 50 and carry significant mortality risk. Current surveillance relies on intermittent imaging (ultrasound or MRI) at 6–24 month intervals, which may miss rapid growth acceleration between visits. We investigate the feasibility of continuous aneurysm diameter tracking using peripheral pulse waves, like those detected by photoplethysmography (PPG) devices.

**Approach:** We use a simplified one-dimensional hemodynamic model that simulates pulse wave propagation from the heart to the pedal digital artery. We first demonstrate diameter estimation when the hemodynamic model parameters defining systemic circulation are known within bounds for an individual, aggregating thousands of observations over hours or days. We then address the more challenging scenario where systemic circulation parameters are only known to be within wider population-level physiological bounds, using a sequential Monte Carlo approach that combines ensemble MCMC with Kalman filtering to marginalise over unknown parameters while tracking the aneurysm diameter. Both approaches are validated through 12-month tracking simulations with constant and accelerating aneurysm growth rates.

**Main results:** While single-observation diameter estimation is fundamentally limited by noise and confounding variables, aggregating 1,600 measurements under baseline noise conditions reduces diameter uncertainty to 0.8 mm when patient-specific hemodynamic parameters are known within bounds. In this setting, tracking simulations across eight virtual patients achieve average root-mean-square error (RMSE) of ∼0.3 mm. When systemic parameters are known only within population-level bounds, joint Bayesian estimation over the full parameter space achieves a median RMSE of 0.65 mm (1.4±0.3 mm, mean±standard error) across 50 virtual patients, remaining within clinically relevant ranges despite the underlying parameters being only partially identifiable.

**Significance:** These physically-grounded, computational results suggest that peripheral pulse wave monitoring through wearable PPG sensors could complement traditional imaging for aneurysm surveillance, potentially enabling earlier detection of growth acceleration and more timely clinical intervention.

## 1 Introduction

Aortic aneurysms represent a significant public health burden, with abdominal aortic aneurysms (AAA) affecting 4–8% of men and 0.5–1.5% of women over age 65 (Kent et al., 2014). These pathological dilations of the aortic wall can remain asymptomatic before sudden rupture, which carries high mortality rates (Reimerink et al., 2013; Vorp and Geest, 2005). Current clinical practice manages aneurysms through periodic imaging surveillance. Guidelines recommend surgery when aneurysm diameter ≥55 mm or when growth rate 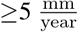(Elefteriades, 2008; Chaikof et al., 2018).

However, imaging is typically performed at 6–24 month intervals using computed tomography (CT), magnetic resonance imaging (MRI), or ultrasound which can underestimate growth rates and risk aneurysm rupture between imaging episodes (Sherifova and Holzapfel, 2019). These limitations motivate investigation of continuous monitoring approaches that could complement traditional imaging. An ideal technology would provide frequent, non-invasive measurements of aneurysm size at home, alerting clinicians to unexpected changes while providing reassurance during stable periods.

Photoplethysmography (PPG) has emerged as a ubiquitous sensing modality in wearable devices and contains rich information about cardiovascular function beyond oxygen saturation (Elgendi, 2012). Features extracted from the PPG waveform—including pulse transit time, augmentation index, and reflection index—have been shown to correlate with arterial stiffness, blood pressure, and vascular health (Millasseau et al., 2006; Charlton et al., 2022a; Karimpour et al., 2023; Charlton et al., 2022b; Alastruey et al., 2023). An aneurysm creates a region of increased compliance and altered impedance within the arterial tree (Humphrey and Holzapfel, 2012), modifying pulse wave propagation from the heart to peripheral measurement sites (Sakalihasan et al., 2018). These modifications may manifest as changes in waveform morphology, timing, and reflection characteristics—all potentially detectable in PPG signals recorded at the periphery (wrist or ankle).

In this work, we present a computational proof-of-concept demonstrating the feasibility of tracking aortic aneurysm diameter using continuous PPG monitoring (Humphrey and Taylor, 2008). Our approach combines a hemodynamic model with Bayesian sequential estimation to aggregate information from thousands of PPG measurements, exploiting natural physiological variation to resolve the diameter signal from confounding variables. Specifically, we make the following contributions:

1. We characterize the fundamental sensitivity and identifiability limitations of single-observation diameter estimation from PPG features, demonstrating that the problem is severely ill-conditioned.
2. When hemodynamic model parameters defining systemic circulation are known within bounds for an individual, we show theoretically via Cramér-Rao bound analysis that aggregating hundreds to thousands of observations— feasible with continuous wearable monitoring—can reduce diameter uncertainty to sub-millimeter levels.
3. We demonstrate accurate 12-month diameter tracking (average RMSE < 0.35 mm) for virtual patients with both constant and accelerating growth rates.
4. We expand the tracking framework and evaluate its robustness when hemodynamic model parameters are not known, showing that clinically acceptable tracking accuracy is maintained despite uncertainties in parameter estimation.

We emphasize that all results presented here are based on a simplified 1D hemodynamic model; the true test of this framework will require expanded high-fidelity computational modeling alongside prospective clinical validation correlating PPG-derived estimates with imaging-confirmed diameter measurements. The gap between idealized simulations and real-world signal quality, patient heterogeneity, and device variability remains substantial. Nonetheless, this work establishes theoretical feasibility, identifies the critical requirements, and presents methodological approaches that future studies can benefit from. We share all data and code to enable full reproducibility and future studies.

## 2 Methods

To establish the theoretical feasibility of estimating AAA diameter from PPG signals, we modeled the arterial pathway as a single, one-dimensional compliant tube extending from the aortic root to the foot. The hemodynamics within this pathway are governed by the nonlinear 1D Navier–Stokes equations—solved via a Lax-Friedrichs scheme—utilizing a Kelvin–Voigt tube law to capture wall mechanics and a three-element Windkessel boundary condition to represent the terminal microcirculation. While simplified, this single-pathway formulation is a well-established approximation in pulse-wave hemodynamics and an appropriate fidelity for feasibility analysis connecting PPG features with 1D hemodynamic biophysical parameters (Sherwin et al., 2003; Alastruey et al., 2012; Van de Vosse and Stergiopulos, 2011; Mynard and Smolich, 2015; Alastruey et al., 2011; Charlton et al., 2019; Willemet and Alastruey, 2015).

### Governing Equations

The model solves the 1D Navier-Stokes equations for blood flow in compliant vessels, consisting of conservation of mass and momentum:

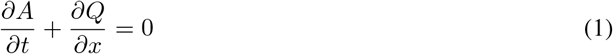

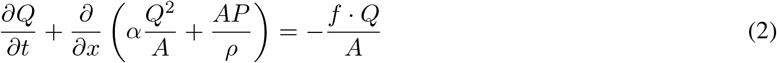

where *A*(*x, t*) is the vessel cross-sectional area [m^2^], *Q*(*x, t*) is the volumetric flow rate [m^3^/s], *P* (*x, t*) is the transmural pressure [Pa], *ρ* = 1060 kg/m^3^ is blood density, *α* = 1.0 is the momentum correction coefficient (flat velocity profile), and *f* represents viscous friction forces.

### Constitutive Relations

Vessel wall mechanics are described by a nonlinear tube law relating pressure to area:

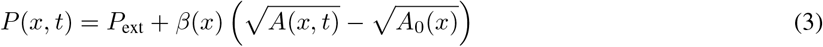

where *P*_ext_ = 0 Pa is external pressure, *A*_0_(*x*) is the reference cross-sectional area at zero transmural pressure, and *β*(*x*) [Pa · m] is the wall stiffness parameter. The local pulse wave velocity is:

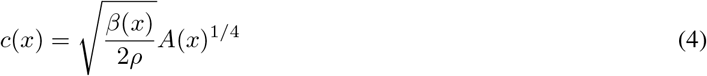

Viscous friction follows a Poiseuille-like profile:

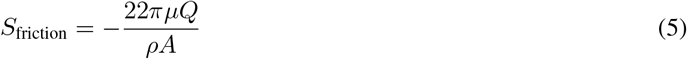

where *µ* = 0.004 Pa · s is blood dynamic viscosity.

### Arterial Geometry and Aneurysm Modeling

The computational domain spans *L* = 1.0 m from the proximal aorta to the pedal digital artery, discretized with spatial step Δ*x* = 0.01 m. The baseline geometry features exponential tapering from proximal radius 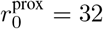 mm to distal radius 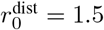mm:

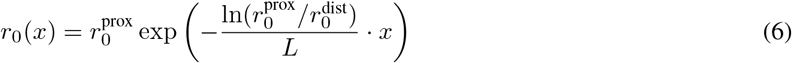

Wall stiffness increases distally to reflect physiological arterial stiffening, ranging from *β*^prox^ = 5 × 10^4^ Pa · m (yielding PWV ≈ 5 m/s in the aorta) to *β*^dist^ = 2 × 10^6^ Pa · m (PWV ≈ 15 m/s in pedal digital arteries).

AAA geometry is incorporated via a Gaussian dilation envelope centered at *x* = 0.35 m (abdominal region) with half-width 5 cm. For a specified maximum diameter *D*_max_, the local area is modified as:

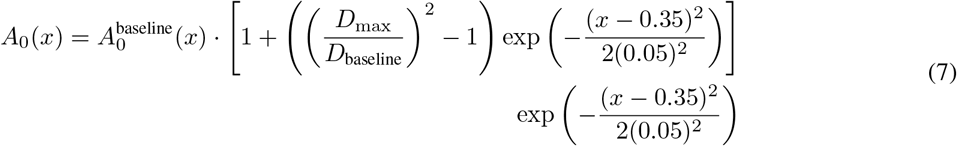

Aneurysmal wall degradation is modeled as a 40% local reduction in *β*(*x*) following the same Gaussian envelope, representing increased local compliance.

### Numerical Method

The system is solved using the Lax-Friedrichs finite difference scheme, chosen for its stability properties. The time step satisfies the Courant-Friedrichs-Lewy (CFL) condition with CFL number 0.4:

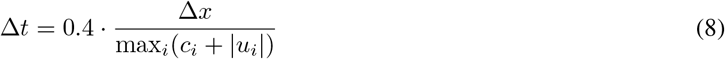

At the inlet (proximal aorta), a physiologically realistic cardiac flow waveform is prescribed, consisting of rapid systolic ejection (35% of cycle), brief early diastolic backflow representing valve closure, and zero diastolic flow. Stroke volume (baseline 70 mL) is adjusted based on mean arterial pressure (MAP) with sensitivity of −1% per mmHg deviation from 90 mmHg baseline.

At the outlet (pedal digital artery), a three-element Windkessel model (*R*_1_-*C*-*R*_2_) represents peripheral impedance, with characteristic impedance *R*_1_ = 5 × 10^7^ Pa · s/m^3^, peripheral resistance *R*_2_ = 5 × 10^8^ Pa · s/m^3^, and compliance *C* = 2 × 10^−9^ m^3^/Pa. Our method generates physiologically realistic peripheral wave and PPG waveforms (Appendix A).

### Feature Extraction

Each simulation runs for 5 cardiac cycles with the final cycle recorded for analysis. Proximal pressure and distal area (PPG proxy) waveforms are extracted. Seven hemodynamic features are computed: 1) Pulse Transit Time (PTT): Foot-to-foot delay between proximal pressure and distal area waveforms [ms], 2) Effective Pulse Wave Velocity (PWV): *L/*PTT [m/s], 3) Augmentation Index (AI): Derived from diastolic decay rate, reflecting wave reflection, 4) Reflection Index (RI): Ratio of diastolic to total pulse area, 5) Stiffness Index (SI): Approximated as effective PWV [m/s], 6) Systolic Time Ratio (STR): Fraction of cardiac cycle in systole, and 7)Dicrotic Notch Timing (DNT): Time from peak to dicrotic notch [ms].

When solving inverse problems (e.g. inferring aneurysm diameter or 1D model parameters), we restrict ourselves to the four features computable from the PPG waveform alone: AI, RI, STR, and DNT.

### Parameters defining systemic circulation

The 8 parameters that characterize systemic circulation 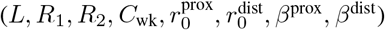 are referred to as *θ*_circ_ and their values as used in our work are provided in the columns of Table 1. We report which values are used and why for each experiment.

**Table 1:** Ranges for Systemic Circulation Parameters (*θ*_circ_) and Usage Across Experiments.

| Parameter | Units | Bounds | Nominal<sup>†</sup> | Nominal ± noise<sup>‡</sup> |
| --- | --- | --- | --- | --- |
| $L$ (Length) | m | [0.7, 1.3] | 0.95 | $\pm 10\%$ |
| $R_1$ (Prox. Resistance) | $\text{Pa} \cdot \text{s}/\text{m}^3$ | $[2 \times 10^7, 1 \times 10^8]$ | $4.47 \times 10^7$ | $\pm 20\%$ |
| $R_2$ (Dist. Resistance) | $\text{Pa} \cdot \text{s}/\text{m}^3$ | $[1 \times 10^8, 1 \times 10^9]$ | $3.16 \times 10^8$ | $\pm 40\%$ |
| $C_{wk}$ (Windkessel Comp.) | $\text{m}^3/\text{Pa}$ | $[5 \times 10^{-10}, 5 \times 10^{-9}]$ | $1.58 \times 10^{-9}$ | $\pm 20\%$ |
| $r_{0\_proximal}$ (Prox. Radius) | m | [0.008, 0.016] | 0.0113 | $\pm 10\%$ |
| $r_{0\_distal}$ (Dist. Radius) | m | [0.001, 0.002] | 0.0014 | $\pm 10\%$ |
| $\beta_{proximal}$ (Prox. Stiffness) | Pa | $[2 \times 10^4, 1 \times 10^5]$ | $4.47 \times 10^4$ | $\pm 25\%$ |
| $\beta_{distal}$ (Dist. Stiffness) | Pa | $[8 \times 10^5, 4 \times 10^6]$ | $1.79 \times 10^6$ | $\pm 25\%$ |

<sup>†</sup> Nominal values are calculated as the geometric mean of the trained physiological bounds: $\theta_{\text{nom}} = \sqrt{\theta_{\text{min}} \cdot \theta_{\text{max}}}$ and used for a representative individual.
<sup>‡</sup> We use these ranges to simulate the biological variance of $\theta_{\text{circ}}$ within a representative individual.

### 2.1 Sensitivity Analysis

To quantify the sensitivity of PPG features to aneurysm diameter, we computed each feature across a physiologically relevant aneurysm diameter range (25–55 mm) while holding constant the heart rate (70 bpm), MAP (95 mmHg), and *θ*_circ_ (Nominal, Table 1). For each feature *f*, we calculated the sensitivity *∂f/∂D* using linear regression over the diameter range. To assess signal-to-noise ratio, we normalized each sensitivity by the corresponding measurement noise standard deviation *σ*_noise_: PTT (*σ* = 2.0 ms), PWV (*σ* = 0.15 m/s), AI (*σ* = 0.03), RI (*σ* = 0.03), SI (*σ* = 0.15 m/s), systolic time ratio (*σ* = 0.03), and dicrotic notch timing (*σ* = 5.0 ms). The normalized sensitivity *∂f*/*∂D* /*σ*_noise_ quantifies how many standard deviations of noise correspond to a 1 mm change in diameter.

### 2.2 Measurement Uncertainty Propagation and Identifiability Analysis

To evaluate how inherent clinical measurement noise in continuously sensed hemodynamics acts as a confounding factor when inferring absolute structural aneurysm diameter, we performed a signal-to-noise sensitivity analysis. Unlike static structural parameters, HR and MAP must be constantly tracked via external sensing devices, each harboring distinct error profiles.

We established a baseline inference scenario centered around a true aneurysm diameter *D* = 45.0 mm, monitored during a nominal physiological state of HR = 75 bpm and MAP = 100 mmHg while *θ*_circ_ was held at Nominal values (Table 1). Measurement errors for HR and MAP were defined at *σ*_HR_ = 1.0 bpm and *σ*_MAP_ = 10.0 mmHg (Appendix B). The extracted peripheral PPG features were subjected to independent, empirically defined measurement errors specific to each scalar (*σ*_AI_ = 0.03, *σ*_RI_ = 0.03, *σ*_STR_ = 0.03, and *σ*_DNT_ = 5.0 ms).

We computed the local Jacobian sensitivity matrix **J** bounding the partial derivatives of each tracked PPG feature with respect to the input diameter, HR, and MAP axes (**J**_*i,j*_ = *∂f*_*i*_/*∂θ*_*j*_). To construct normalized Signal-to-Noise Ratios (SNR), each element of **J** was divided by the corresponding feature’s operational measurement standard deviation. The conditioning of the resultant noise-normalized Jacobian was analyzed with Singular Value Decomposition (SVD) to quantify the degree to which observational HR and MAP uncertainties effect diameter estimates over a single continuous measurement frame. The condition number *κ*(**J**) quantifies parameter correlation: *κ* < 10 indicates excellent identifiability, 10 ≤ *κ* < 100 moderate identifiability, 100 ≤ *κ* < 1000 poor identifiability, and *κ* ≥ 1000 very poor identifiability.

In a unified visualization, 2D feature response heatmaps were calculated across the dense operating grid (*D* ∈ [25, 55] mm, HR ∈ [50, 120] bpm, MAP ∈ [55, 180] mmHg, *θ*_circ_ held constant at Nominal values, Table 1), overlaid with explicit probability ellipses (2*σ*) corresponding to the defined clinical error covariances.

### 2.3 Cramér-Rao Bound Analysis for Aneurysm Diameter

To quantify how repeated longitudinal measurements reduce uncertainty in estimating the aneurysm diameter *D*, we performed an extended Cramér-Rao Bound analysis that accounts for natural physiological variation in hemodynamics. While continuous monitoring naturally yields many samples, intra-patient variations in systemic properties present moving operating points. We simulated a continuous monitoring scenario where the true aneurysm diameter was fixed (*D* = 45.0) over an observation window (e.g., up to 24 hours) with varying physiological state (HR ∼ 𝒰 (60, 90) bpm, MAP ∼ 𝒰 (80, 110) mmHg, and *θ*_circ_ ∼ Nominal ± noise).

The Fisher information for diameter was calculated as:

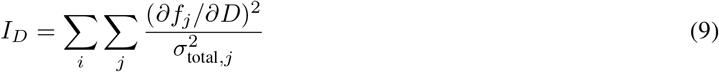

where the sum over *i* represents multiple measurements with different HR and MAP values, the sum over *j* represents the PPG-only features, and 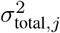accounts for both intrinsic feature noise and propagated uncertainty from HR/MAP measurements:

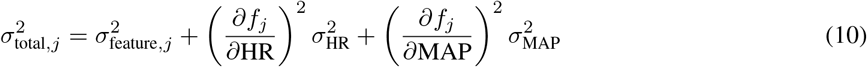

The CRLB on diameter uncertainty is 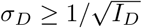We tested four noise multipliers (0.5, 1, 2, 4) applied to baseline uncertainties (*σ*_HR_ = 1 bpm, *σ*_MAP_ = 10 mmHg, *σ*_AI_ = 0.03, *σ*_RI_ = 0.03, *σ*_STR_ = 0.03, and *σ*_DNT_ = 5.0 ms) and three measurement counts (*N* = 400, 1600, 6400 observations). Each measurement represents 5 cardiac cycles with the tested measurement counts ranging from ∼0.5–8.0 hours.

### 2.4 Neural Surrogate Model

To enable computationally tractable inverse problems, we developed a neural network surrogate approximating the 1D Navier-Stokes solver. The surrogate is a fully connected multilayer perceptron (MLP) with 11-dimensional input comprising: aneurysm diameter *D* (mm), heart rate HR (bpm), MAP (mmHg), vessel length *L* (m), proximal resistance *R*_1_ (Pa s/m^3^), distal resistance *R*_2_ (Pa s/m^3^), Windkessel compliance *C*_wk_ (m^3^/Pa), proximal radius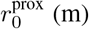, distal radius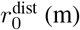, proximal stiffness *β*^prox^ (Pa), and distal stiffness *β*^dist^ (Pa). The network has three hidden layers with [128, 128, 64] neurons using ReLU activations and dropout (*p* = 0.1). The 7-dimensional output predicts PTT, PWV_eff_, AI, RI, SI, systolic time ratio, and dicrotic notch timing. The network contains approximately 30,000 trainable parameters.

Training data was generated using the 1D hemodynamic model with Latin Hypercube Sampling over the 11-dimensional parameter space, with 40,000 samples for models where parameter ranges were representative of the population (*D ∈* [25, 55] mm, HR ∈ [50, 120] bpm, MAP [55, 180] mmHg, and *θ*_circ_ ∈Bounds, Table 1). The network was trained with Adam optimizer (learning rate *α* = 0.001), MSE loss, batch size 64, and early stopping (patience 10 epochs) monitoring validation loss. On held-out validation sets, the surrogate achieved *R*^2^ > 0.88 for all features, with relative errors < 5% for most features (Appendix C). The surrogate provides approximately 1000 × speedup compared to the numerical solver and is used to approximate the 1D model when solving inverse problems (Pant et al., 2014).

### 2.5 Bayesian Sequential Diameter Tracking with Known *θ*_circ_

We implemented a Kalman-style, Bayesian tracking framework that fuses biomechanical growth model priors with PPG-based diameter observations. The prior distribution at time *t* is computed from the posterior at *t* − Δ*t* propagated forward:

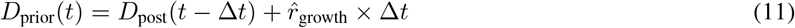

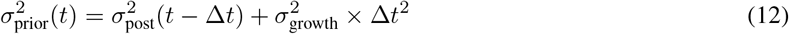

where 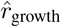 is the adaptively estimated growth rate and *σ*_growth_ = 0.3 mm/month represents growth model uncertainty.

At each timepoint, synthetic PPG observations are generated with the 1D hemodynamic model by computing true PPG features from true diameter, true HR, true MAP, and *θ*_circ_ sampled uniformly from Nominal±noise (Table 1). Each observation was also adjusted by adding appropriately scaled measurement noise (*σ*_HR_ = 1 bpm, *σ*_MAP_ = 10 mmHg, *σ*_AI_ = 0.03, *σ*_RI_ = 0.03, *σ*_STR_ = 0.03, and *σ*_DNT_ = 5.0 ms). We aggregate 1000s of simulated, noisy observations for a single measurement event where we invert the hemodynamic model to find the current aneurysm diameter from the synthetic observations. The inversion is formulated as an optimization problem solving for aneurysm diameter with the SLSQP algorithm using the neural surrogate for tractability and solved within a local search window around the prior diameter estimate (± 5 mm). During inversion, we use a point-based estimate of *θ*_circ_ at the Nominal values (Table 1). Observation uncertainty was set to *σ*_PPG_ = 1.0 mm.

The posterior distribution is obtained via Bayesian fusion:

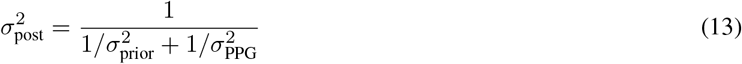

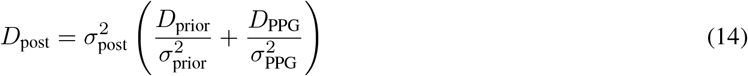

The growth rate estimate is updated using exponential moving average with learning rate *α* = 0.3:

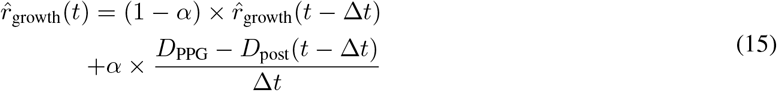

#### 2.5.1 Tracking Simulation Protocol

We simulated eight virtual patients over 12 months with four measurement events per month (48 total). The cohort included four patients with constant growth rates (initial diameters 30, 35, 40, 45 mm with growth rates 0.1, 0.1, 0.2, 0.5 mm/month) and four with accelerating growth (same initial diameters, zero growth for months 0–6, then matching rates for months 6–12) (Sweeting et al., 2012).

Each measurement comprised 3,200 observations generated by the forward 1D-hemodynamic model (HR∼ 𝒰 (60, 90) bpm, MAP ∼ 𝒰 (80, 110) mmHg, and *θ*_circ_∼ Nominal ± noise). The Bayesian tracker (Section 2.5) had uncertain knowledge of initial growth rate and no knowledge of future acceleration, relying on HR, MAP, and PPG observations to detect changes.

### 2.6 Diameter Tracking with Unknown Systemic Circulation Parameters

The experiments presented so far assumed *θ*_circ_ was known at Nominal values or varied within Nominal±noise (Table 1), representing an individual whose systemic circulation had been characterised to within biologically realistic intra-individual variance. We now ask whether aneurysm diameter tracking remains tractable when *θ*_circ_ is known only to lie within the full population-level physiological bounds (Table 1).

We proceed in two stages. First, we compute the Fisher Information Matrix (FIM) and Cramér-Rao Bounds (CRB) for *θ*_circ_ to establish the theoretical identifiability limits of these parameters from PPG features alone. Second, we combine ensemble MCMC sampling with the Bayesian sequential tracking framework of Section 2.5 to jointly estimate *θ*_circ_ and *D* over time, using the neural surrogate for computational tractability. This follows methods similar to those used by others for solving inverse problems in blood flow modeling (Nolte and Bertoglio, 2022; Bertoglio et al., 2012; Pant et al., 2014; Arthurs et al., 2020; Lal et al., 2017).

#### 2.6.1 FIM and Cramér-Rao Bounds for *θ*_circ_

Let *θ*_circ_ ∈ ℝ^8^ denote the systemic circulation parameter vector and let **f**_*i*_ = *g*(*θ*_circ_, *D*_*i*_, HR_*i*_, MAP_*i*_) + ***ϵ***_*i*_ be the observation model for the *i*-th measurement, where *g*(.) is the trained surrogate forward model and ***ϵ***_*i*_ *∼* 𝒩 (**0, Σ**_*ϵ*_) is additive measurement noise with diagonal covariance **Σ**_*ϵ*_ parameterised by the clinical feature variances defined in Section 2.3.

For *N* physiologically independent measurements the FIM is:

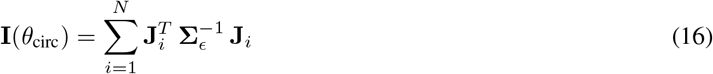

where **J**_*i*_ = *∂g*/*∂θ*_circ_ ∈ ℝ^7×8^ is the Jacobian evaluated at the *i*-th measurement conditions via central finite differences. The CRB on the variance of any unbiased estimator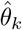 is:

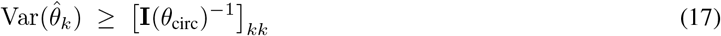

We performed three complementary analyses:

1. **FIM eigen-structure**. Eigenvalue decomposition of the FIM at the Nominal operating point (Table 1) with *N* = 100 measurements, quantifying the conditioning of the inverse problem.
2. **Measurement scaling**. Relative CRB (*σ*_CRB_*/θ*) as a function of *N* ∈ {10, 100, 500, 1000, 5000, 10, 000} under noise multipliers of 0.5×, 1.0×, and 2.0×.
3. **Population variability**. Because local identifiability varies across the nonlinear parameter space, we repeated the CRB analysis for 100 fictive patients whose *θ*_circ_ values were sampled uniformly from the population bounds (Table 1), evaluating identifiability at *N* = 100, 1, 000, and 5, 000 measurements per patient.

#### 2.6.2 Joint Posterior for Diameter and Systemic Parameters

At each time step *t* the patient generates *N* noisy PPG measurements **f**_*t*_ = *f*_*t*,1_, …, *f*_*t,N*_, all produced at the same true diameter *D*_*t*_. The joint log-posterior is:

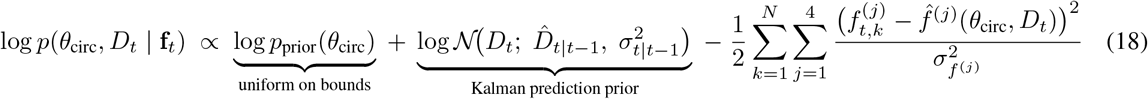

where 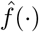denotes the surrogate prediction and 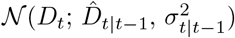is the Kalman prediction prior from Section 2.6.5. Crucially, the likelihood sums over the *current month’s measurements only*; information about *θ*_circ_ from earlier months is carried forward through the MCMC walker positions, as described next.

#### 2.6.3 Sequential MCMC with Warm-Starting

We sample the joint posterior (Eq. 18) using the affine-invariant ensemble sampler emcee (Foreman-Mackey et al., 2013) with 128 walkers. At each subsequent time step (*t >* 1), the ensemble is *warm-started* from the previous month’s final walker positions. A small jitter is applied: 0.5% multiplicative Gaussian noise on *θ*_circ_ (whose true values are constant) and additive noise scaled by *σ*_growth_ Δ*t* on *D*_*t*_ (which changes with growth). This yields the sequential Bayesian update:

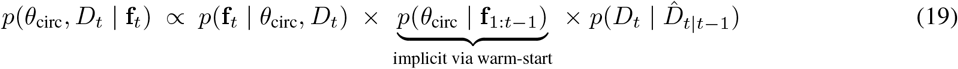

Because the walkers already reside in the previous posterior, no explicit parametric approximation of *p*(*θ*_circ_ **f**_1:*t*−1_) is required. Parameter uncertainty narrows organically as longitudinal data accumulates, without the bias that would arise from fixing *θ*_circ_ at a point estimate.

#### 2.6.4 Initialization via Log-Space Optimization

At *t* = 1 the walkers must be initialised without prior samples. Uniform sampling of the 9-dimensional space is inefficient because the parameter scales span 17 orders of magnitude (*C*_wk_ ∼ 10^−9^ vs. *R*_2_ ∼ 10^8^). We therefore perform 128 multi-start L-BFGS-B optimizations of the negative log-posterior with the *θ*_circ_ components parameterised in log-space, while *D* remains in natural units. The walkers are then distributed around the top modes with small Gaussian perturbations, providing a well-concentrated initialisation near the posterior’s typical set.

#### 2.6.5 Kalman Fusion and Growth Rate Estimation

After discarding burn-in samples, we extract the marginal posterior mean *µ*_ppg_ and standard deviation *σ*_ppg_ of *D*_*t*_ from the MCMC chain. These serve as the observation (*D*_PPG_, *σ*_PPG_) in the Kalman fusion step of Section 2.5, replacing the optimization-based diameter estimate used in the known-*θ*_circ_ case. The Kalman predict, fuse, and growth-rate update (Equations 11–15) are otherwise identical. The fused posterior diameter then seeds the Kalman prediction prior for the next month’s MCMC run, closing the sequential loop.

#### 2.6.6 Virtual Patient Cohort and Tracking Protocol

In the known-*θ*_circ_ experiments of Section 2.5.1, all eight virtual patients shared the same Nominal baseline for their systemic circulation parameters, with intra-individual physiological variation modeled by sampling *θ*_circ_ ∼ Nominal ± noise (Table 1) at each measurement event. This represents a scenario in which a patient’s systemic circulation has been characterized—for example, through an extensive initial imaging-calibrated session—and only beat-to-beat biological variability remains.

Here, we model a more realistic clinical setting in which no patient-specific calibration of *θ*_circ_ has been performed (Charlton et al., 2019). We generated a cohort of 50 virtual patients as follows. For each patient, a baseline 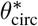was drawn uniformly from the full population-level bounds (Table 1), producing a diverse cohort spanning the physiological range. During tracking, each measurement event sampled *θ*_circ_ from 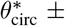 noise, using the same relative noise percentages as in Section 2.5.1 (Table 1). This two-level sampling scheme separates inter-patient diversity (different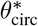 across patients) from intra-patient biological variability (measurement-to-measurement jitter around each patient’s 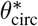).

Crucially, the MCMC-Kalman tracker has no knowledge of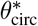; it is given only the population-level bounds as a uniform prior. All other aspects of the simulation—growth rate assignment, measurement generation, and Kalman fusion—follow the same protocol as Section 2.5.1. Initial diameters were drawn uniformly from *D*_0_ ∈[30, 50] mm, growth rates were sampled from {0.03, 0.0625, 0.125, 0.25, 0.5} mm/month, and half the cohort was assigned an acceleration event at month 6. Each monthly measurement event comprised 3,200 observations generated by the forward hemodynamic model under physiologically varying HR, MAP, and *θ*_circ_.

## 3 Results

### 3.1 Model Validation

We first validate that the 1D hemodynamic model produces physiologically realistic PPG-like waveforms at the pedal digital artery (Appendix A). The simulated waveforms exhibit characteristic features including rapid systolic upstroke, dicrotic notch, and diastolic decay. Varying mean arterial pressure across a wide range (30–200 mmHg) produces expected changes in waveform morphology and amplitude, validating the model’s adequate fidelity.

### 3.2 Sensitivity of PPG Features to Aneurysm Diameter

Figure 1 shows the relationship between PPG-derived features and aneurysm diameter across the clinically relevant range (25–55 mm). All features exhibit detectable but modest sensitivity to diameter changes. Pulse wave velocity and stiffness index show monotonically increasing relationships with diameter, while augmentation index and reflection index display more complex, non-monotonic patterns. The normalized sensitivity panel (bottom right) reveals that SI and PWV have the highest signal-to-noise ratios (*∂f*/*∂D* /*σ*_noise_ ≈ 0.05), while dicrotic notch timing provides the most informative PPG-only feature.

**Figure 1:**
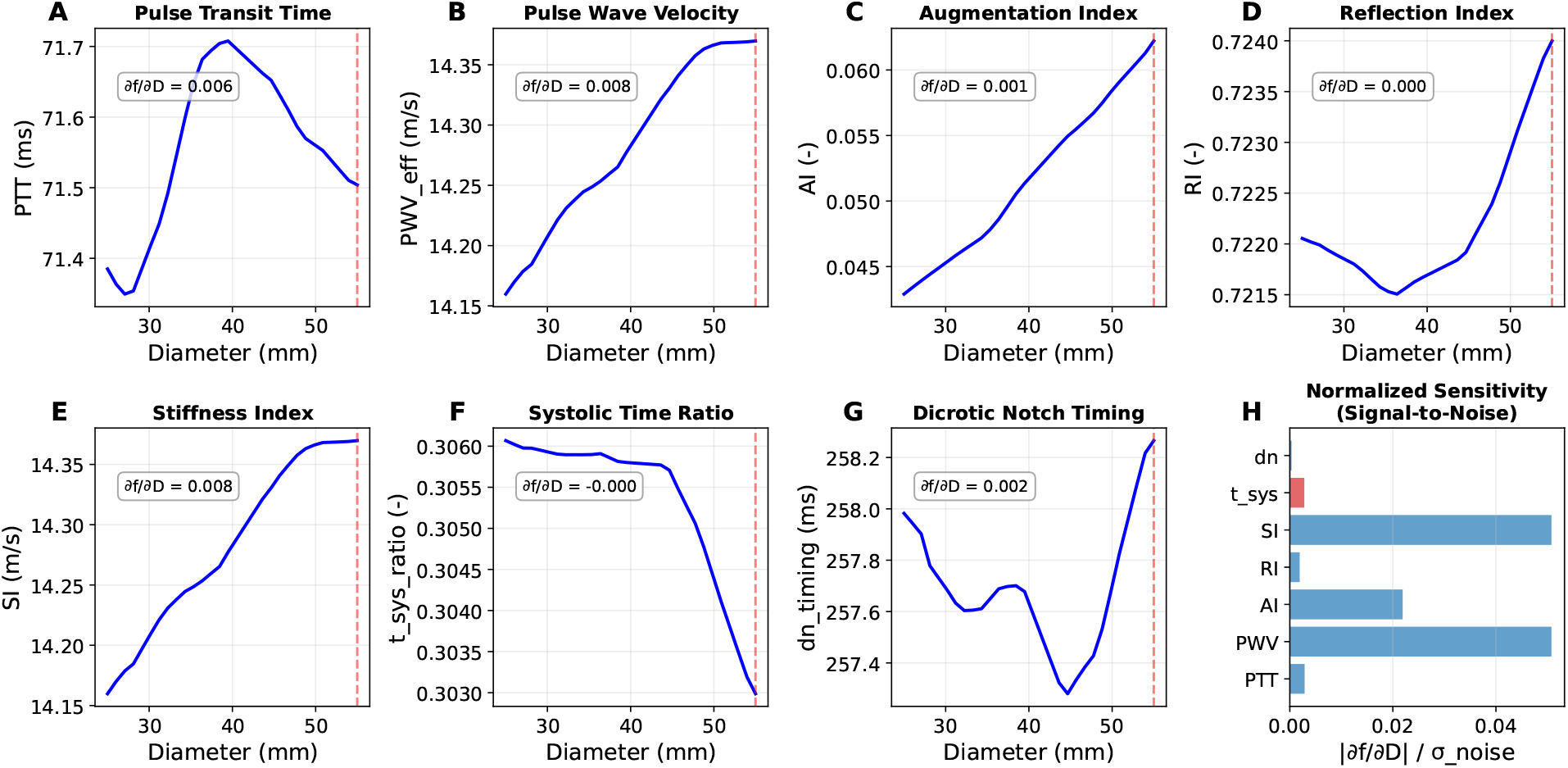
Sensitivity of PPG features to aneurysm diameter. Top row: Pulse transit time, pulse wave velocity, augmentation index, and reflection index versus diameter. Bottom row: Stiffness index, systolic time ratio, dicrotic notch timing, and normalized sensitivity comparison. Vertical dashed lines indicate the 55 mm surgical threshold. Sensitivities (*∂f*/*∂D*) are shown in each panel. The normalized sensitivity panel demonstrates that all features have low signal-to-noise ratios for single-observation diameter estimation.

Critically, these sensitivities are small relative to measurement noise—a 1 mm diameter change produces feature changes of only 0.01–0.05 standard deviations. This fundamental limitation motivates the multi-observation aggregation strategy developed in subsequent sections.

### 3.3 Measurement Uncertainty and Parameter Identifiability

The uncertainty propagation analysis revealed that independent structural diameter estimation from a single measurement point is profoundly ill-posed due to severe confounding with the required HR and MAP inputs. Evaluating the noise-normalized Jacobian at the nominal baseline state (*D* = 45.0 mm, HR= 75 bpm, MAP= 100 mmHg) generated a scaled condition number of *κ* = 739.32. The spread across the singular values (*λ*_1_ = 78.73, *λ*_2_ = 5.02, *λ*_3_ = 0.11) signifies that certain directions in the parameter space are nearly unobservable. Specifically, the analysis verified that perturbations in the true structural diameter *D* produce highly comparable PPG features relative to routine shifts in HR or MAP.

Figure 2 presents the feature space structure and uncertainty propagation analysis. Tracking the 2*σ*-covariance error ellipses projected across the 2D feature heatmaps (e.g., *D* vs. HR and *D* vs. MAP) distinctly highlighted oblique, off-axis stretching. The tilted axes of the uncertainty ellipses trace identically along the physiological feature isoclines. This indicates parameter coupling: if a sensor artificially measures HR slightly higher than truth, the inverse solver will natively compensate by predicting a highly erroneous aneurysm diameter to minimize the forward feature discrepancy. Consequently, these results suggest that single-point estimation of aneurysm diameter from peripheral pulse wave features is essentially impossible and achieving clinical-grade bounds mandates aggregation of measurements.

**Figure 2:**
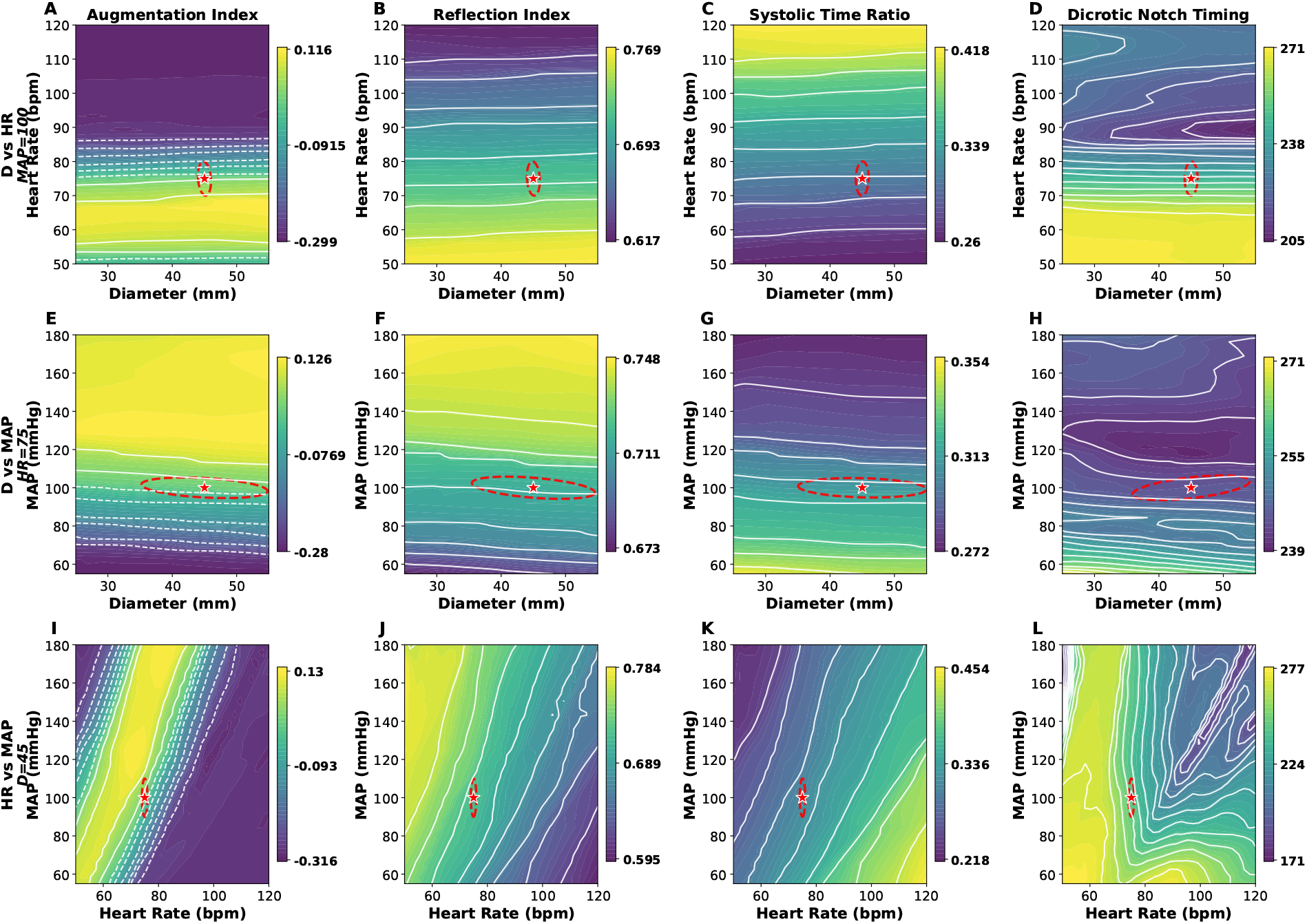
Feature space structure and measurement uncertainty propagation. Rows show cross-sections of the 3D parameter space: diameter vs. heart rate (top), diameter vs. MAP (middle), and heart rate vs. MAP (bottom). Columns correspond to four PPG-only features. Red stars indicate the nominal operating point (D=45 mm, HR=75 bpm, MAP=100 mmHg). Red dashed ellipses show 2*σ* uncertainty regions from HR (heart) and MAP (mean arterial pressure) measurement errors. The elongated ellipses along diameter axes indicate poor single-observation identifiability (condition number *κ* ≈ 740).

### 3.4 Cramér-Rao Bounds for Multi-Observation Diameter Estimation

Figure 3 demonstrates how diameter estimation uncertainty decreases with increasing numbers of observations and varies with measurement noise levels. The left panel shows that at baseline noise (1×), uncertainty decreases from approximately 2.8 mm with 400 measurements to 0.7 mm with 6,400 measurements. Even with elevated noise (4×), 6,400 measurements achieve approximately 1.0 mm precision.

**Figure 3:**
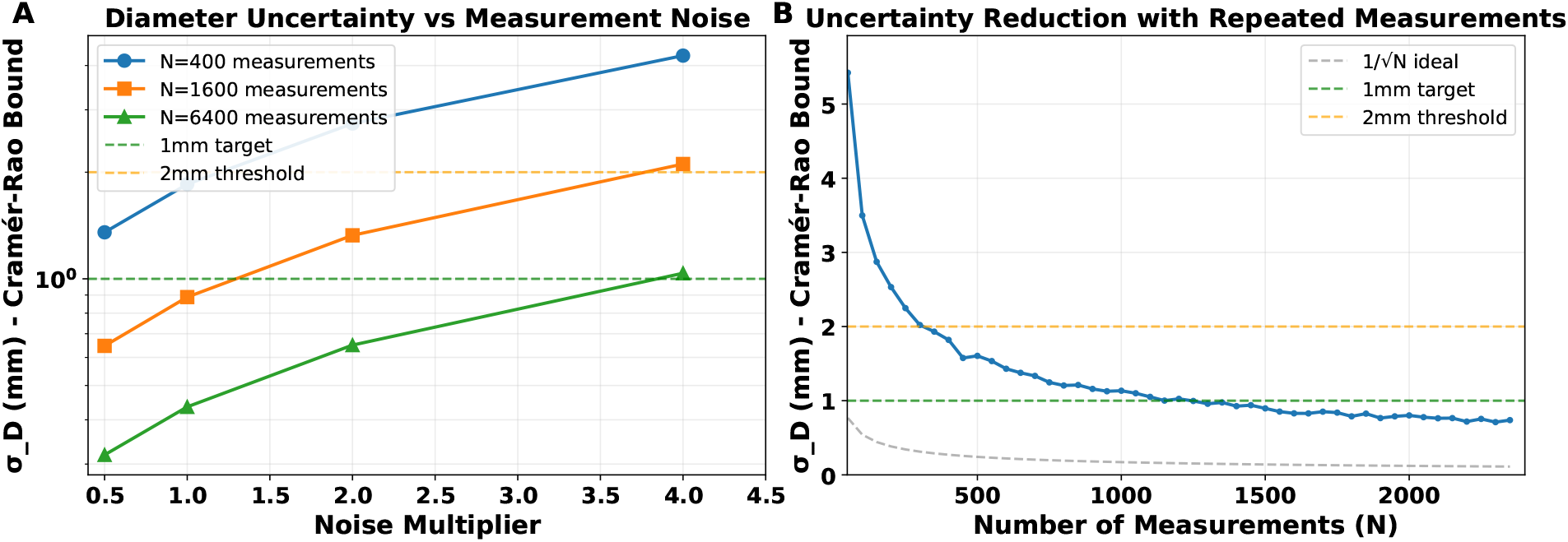
Cramér-Rao lower bounds for diameter estimation. Left: Diameter uncertainty versus noise multiplier for different measurement counts. The 1 mm target (green dashed) and 2 mm clinical threshold (orange dashed) are shown for reference. Right: Uncertainty reduction with increasing measurements at baseline noise, compared to ideal 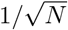 scaling. Sub-millimeter precision is theoretically achievable with 1,600+ measurements.

The right panel in Figure 3 illustrates uncertainty reduction as a function of measurement count at baseline noise. The observed scaling approaches but does not perfectly match the ideal 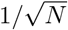 behavior (gray dashed line), reflecting the additional variance contributed by HR and MAP measurement uncertainty. Nonetheless, the results demonstrate that sub-millimeter precision—adequate for clinical tracking—is theoretically achievable with feasible observation counts. Collecting 1,600 observations (approximately 100 minutes of monitoring at 5 cardiac cycles per observation) brings uncertainty below the 1 mm target threshold.

### 3.5 Longitudinal Diameter Tracking with Known *θ*_circ_

Figure 4 presents 12-month tracking results for eight virtual patients as described in Section 2.5.1. The top row shows four patients with constant growth rates; the bottom row shows four patients whose growth accelerated at month 6 (Sweeting et al., 2012).

**Figure 4:**
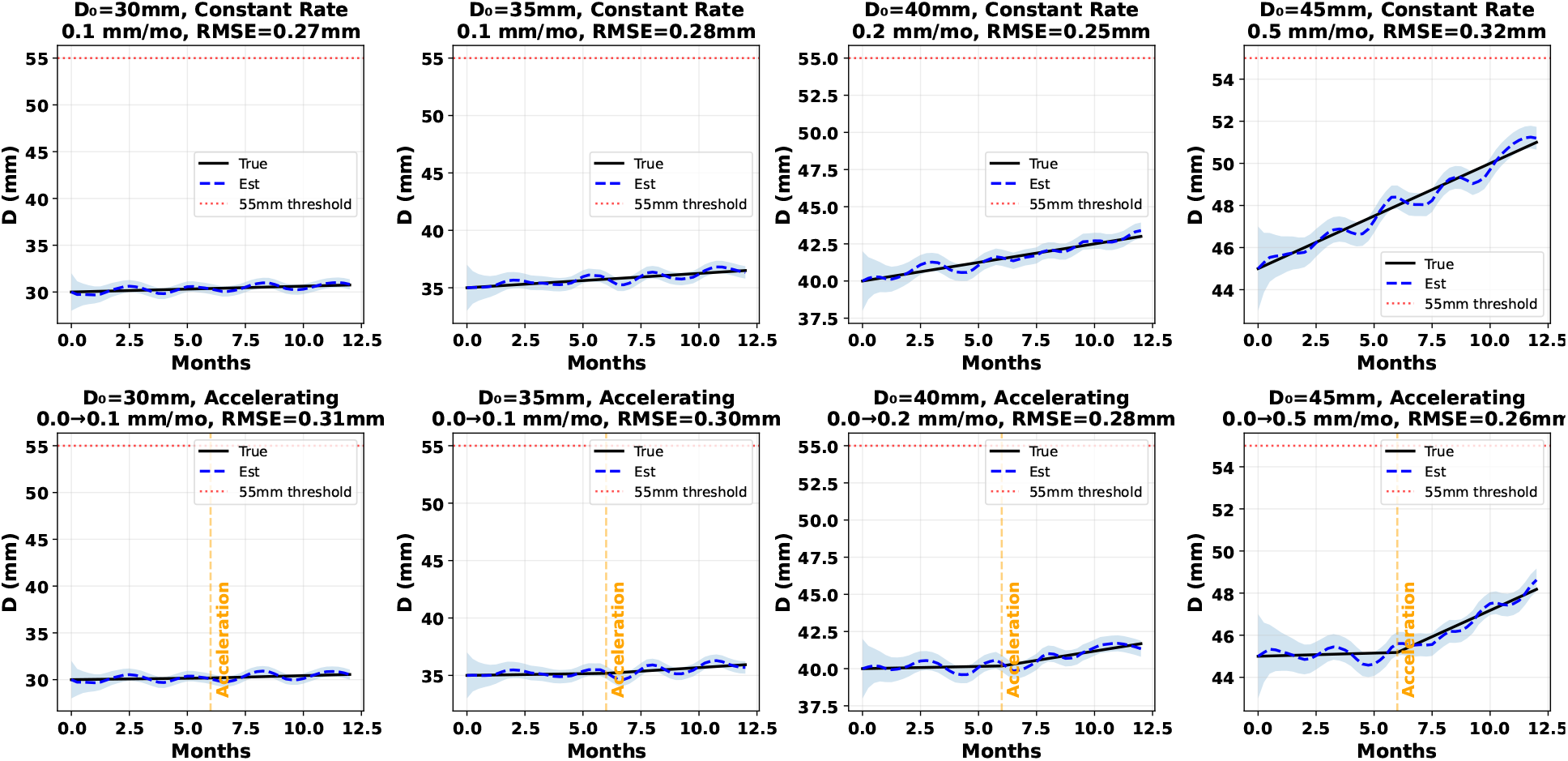
Twelve-month diameter tracking for eight virtual patients as described in Section 2.5.1. Top row: Constant growth rates (0.1, 0.1, 0.25, 0.5 mm/month for initial diameters 30, 35, 40, 45 mm). Bottom row: Accelerating growth (zero growth months 0–6, then matching rates). Black solid lines: true diameter; blue dashed lines: estimated diameter; blue shading: ±2*σ* uncertainty; orange vertical lines: acceleration timepoints; red dotted lines: 55 mm surgical threshold. RMSE values are shown in each panel title.

For constant growth patients, the Bayesian tracker maintains close agreement with true diameter throughout the monitoring period, with RMSE ranging from 0.25–0.32 mm and maximum errors below 0.61 mm. The ±2*σ* uncertainty bands (blue shading) appropriately capture the true diameter trajectory.

For accelerating growth patients, the tracker successfully detects and adapts to the growth rate change despite having no prior knowledge of the acceleration event. Following the acceleration at month 6 (orange vertical lines), the adaptive growth rate estimator increases 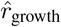 within 1–2 measurement cycles, enabling continued accurate tracking. RMSE values (0.26–0.31 mm) remain comparable to the constant growth cases, demonstrating robustness to dynamic growth patterns.

Across both constant and accelerating growth cohorts the average RMSE was 0.28±0.01 (mean±standard error). This suggests that aneurysm diameter tracking from peripheral pulse waveform features is feasible with clinically-relevant error rates when systemic circulation parameters are known to be within biologically realistic variance from Nominal values (Nominal±noise, Table 1).

### 3.6 Theoretical Parameter Identifiability Limits for Systemic Circulation Parameters (*θ*_circ_)

At the Nominal *θ*_circ_ values with *N* = 100 measurements, the FIM yielded an extreme condition number of 2.23×10^30^ (details in Appendix D). The eigenvalue spectrum spanned numerous orders of magnitude (*λ*_min_ = −4.18×10^4^, *λ*_max_ = 1.21×10^21^) suggesting that features possess highly varying degrees of sensitivity depending on the specific parameter. Correlation analysis revealed strong coupling of estimation uncertainty between parameter pairs [*L,β*_proximal_], [*R*_1_,*R*_2_], and [*β*_proximal_,*β*_distal_] (Appendix D). Together these results suggest that estimating certain linear combinations of parameters from PPG features alone may require large sample sizes and proper posterior estimation (not point-based estimates).

Despite the poor local conditioning at Nominal *θ*_circ_ values, calculating the theoretical limits via FIM inversion showed that all 8 parameters achieve a relative theoretical uncertainty bound (*σ*_CRB_*/θ*) of strictly less than 10% with increasing number of samples. Varying the number of available measurements produced expected convergence rates. With nominal noise (1.0 ×), increasing the batch size from *N* = 10 to *N* = 100 strictly pushed the relative bound for distal resistance *R*_2_ (the most difficult parameter to identify) from 27.6% down to 9.4%. Escalating the noise profile by a factor of 2 predictably degraded performance bounds by roughly a factor of two globally, shifting optimal estimation thresholds such that identifying *R*_2_ beneath a 10% margin required *N* ≥ 500 measurements under high-noise conditions.

The population sweep simulation of 100 distinct fictive patients demonstrated that these identifiability characteristics hold stably across the broader physiological distribution, not just at the Nominal values (Figure 5, boxplot). The relative theoretical uncertainty (*σ*_CRB_*/θ*) for *R*_1_, *R*_2_, and *C*_*wk*_ was generally higher than for other parameters across different numbers of measurements (*N* = 100, 1000, 5000). All parameters could be estimated with *σ*_CRB_*/θ* ≤ 10^−1^ for *N* ≥ 1000.

**Figure 5:**
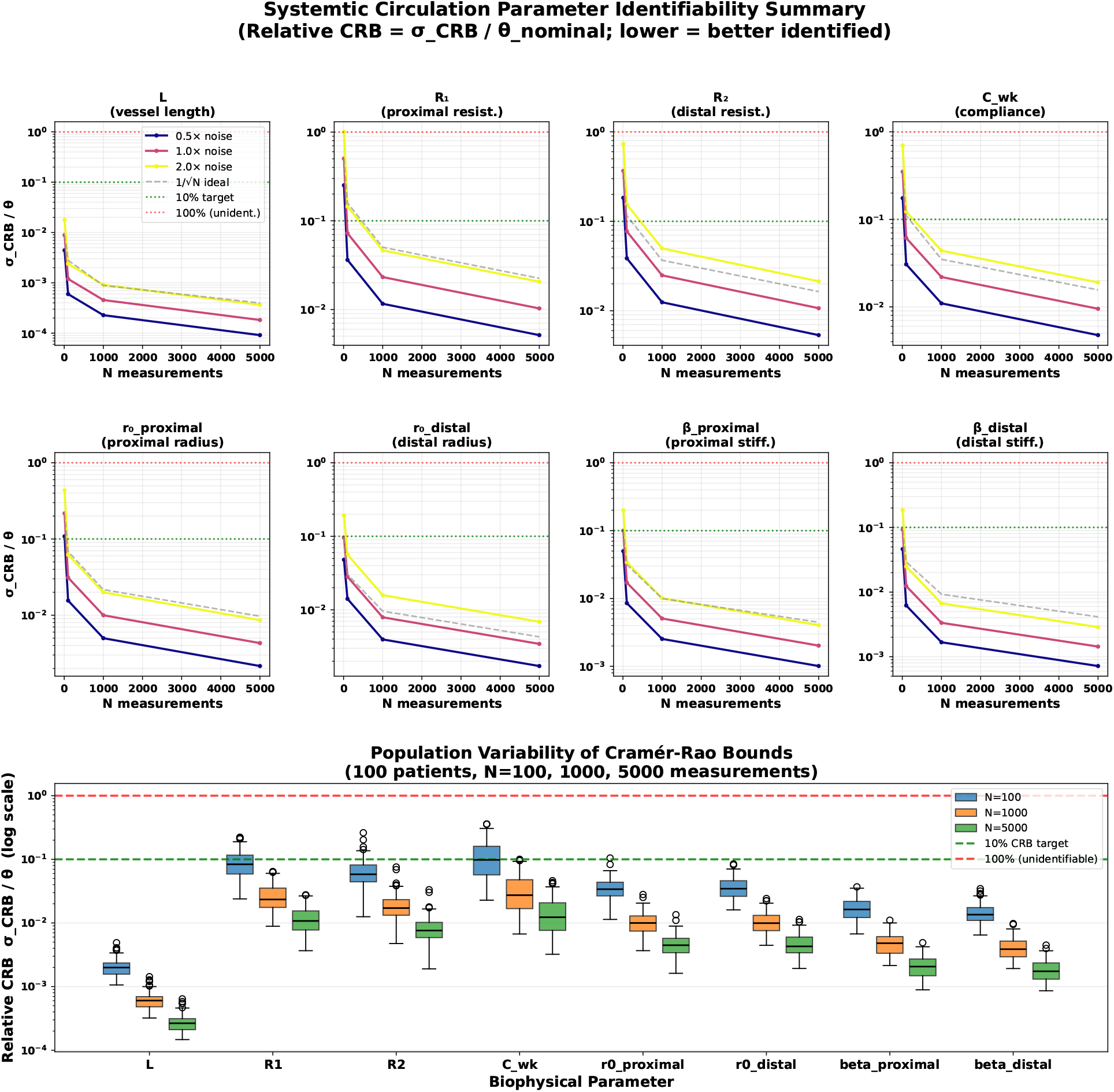
Cramér-Rao Bound (CRB) identifiability analysis for the 1D hemodynamic model’s system circulation parameters.(Top) The relative CRB (*σ*_CRB_*/θ*) as a function of the number of PPG measurements (*N*) for *θ*_circ_, evaluated at nominal operating conditions. The bounds are shown under varying noise levels: baseline feature noise (1.0x, pink), reduced noise (0.5x, blue), and elevated noise (2.0x, yellow). The grey dashed line illustrates the ideal theoretical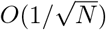 scaling. A target identifiability threshold of 10% relative error (green dotted line) and an unidentifiability limit of 100% relative error (red dotted line) are provided for reference. (Bottom) Population variability of parameter identifiability. Box plots represent the distribution of relative CRBs across a fictive population of 100 patients with randomly sampled system circulation properties within physiological bounds. The distributions are shown for measurement budgets of *N* = 100 (blue), *N* = 1000 (orange), and *N* = 5000 (green). While a budget of *N* = 100 measurements yields median CRBs approaching or exceeding the 10% target for more difficult-to-estimate parameters (e.g., *R*_1_, *C*_*wk*_), increasing the observational budget to *N* 1000 measurements guarantees robust theoretical identifiability (*σ*_CRB_*/θ <* 0.1) for nearly all parameters across the diverse patient population.

However, we emphasize that the CRB is a *lower bound* on variance for unbiased estimators evaluated at a single mode; it does not account for multimodality, surrogate approximation error, or finite MCMC mixing. The CRB results therefore represent a best-case theoretical benchmark against which the practical MCMC tracking performance should be compared.

### 3.7 Tracking Aneurysm Diameter with Sequential MCMC with Kalman fusion with Unknown *θ*_circ_

Across 50 virtual patients, the MCMC-Kalman tracker achieved a mean RMSE of 1.41 ± 0.3 mm, a mean absolute error (MAE) of 1.26±0.3 mm, and a maximum tracking error of 2.01±0.5 mm (mean±standard error). The median RMSE was much lower at 0.65 mm; 3 patients exhibited catastrophic failure (> 6 mm), likely due to issues with parameter identifiability and strong coupling of parameter uncertainty. Representative per-patient tracking trajectories are shown in Figure 6, illustrating the range of behaviors across patients with varying growth rates and growth patterns. The majority of patients exhibit tracking errors well below 1 mm, with the posterior uncertainty bands (shaded region) reliably enclosing the true diameter trajectory. Higher errors are observed in a small subset of cases where growth rate changes occur abruptly mid-trajectory or where systemic circulation parameter identifiability is limited.

**Figure 6:**
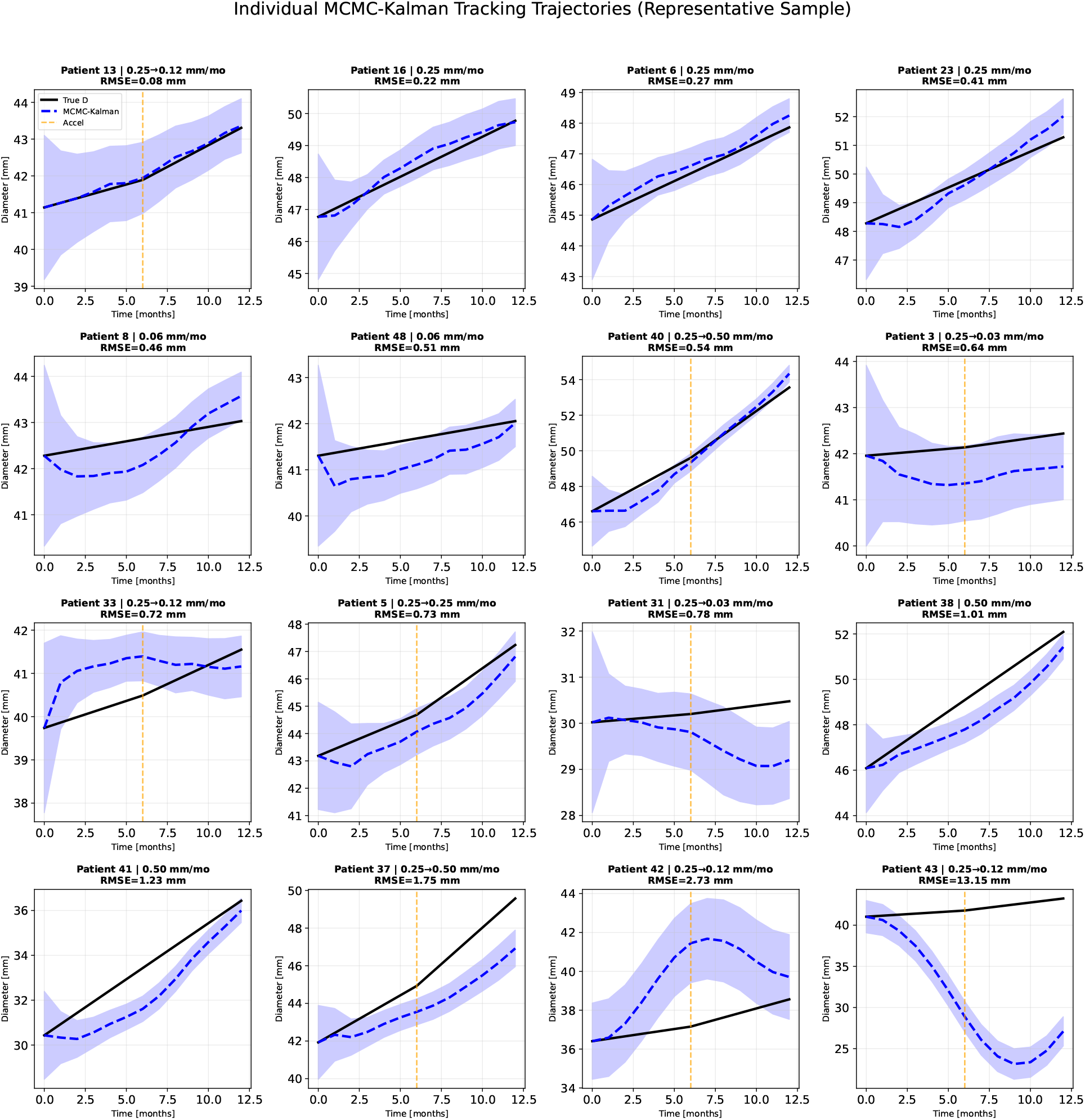
MCMC-Kalman tracking trajectories for a representative cohort of 16 simulated patients with unknown *θ*_circ_ values (Bounds, Table 1). Each panel shows the ground-truth aneurysm diameter (solid black), the MCMC-Kalman posterior mean estimate (dashed blue), and the 95% credible interval (shaded blue), plotted over a 12-month follow-up window. Patients are ordered from lowest to highest RMSE (left-to-right, top-to-bottom), sampled at evenly-spaced percentiles of the full cohort RMSE distribution to span the range of tracking difficulty. Each panel header reports the patient index, growth rate profile, and RMSE. For patients with accelerating growth (rate change at 6 months, e.g. 0.25 → 0.50 mm/mo), a vertical dashed orange line marks the acceleration onset. The tracker achieves sub-millimeter RMSE for the majority of patients, with larger errors concentrated in cases exhibiting rapid or decelerating growth and in one outlier (Patient 43, RMSE = 13.15 mm) where the posterior drifts far from the true trajectory. The average RMSE across all 50 virtual patients was 1.4±0.3 mm (standard error).

MCMC convergence diagnostics, shown in Figure 7, confirm the overall stability of the sampling procedure. The median acceptance fraction across patients remained in the range 0.35–0.45 throughout the 12-month tracking window, consistent with the target range for ensemble samplers, and was stable over time. The Kalman fusion weight assigned to the MCMC observation averaged approximately 0.50, indicating that the MCMC posterior and the Kalman kinematic prediction contributed roughly equally to the fused state estimate. Crucially, both acceptance fraction (*r* = −0.61, *p* < 0.001) and MCMC innovation weight (*r* = −0.63, *p* < 0.001) were significantly negatively correlated with per-patient tracking RMSE, suggesting that low acceptance rates and innovation fractions are a reliable early indicator of tracking difficulty.

**Figure 7:**
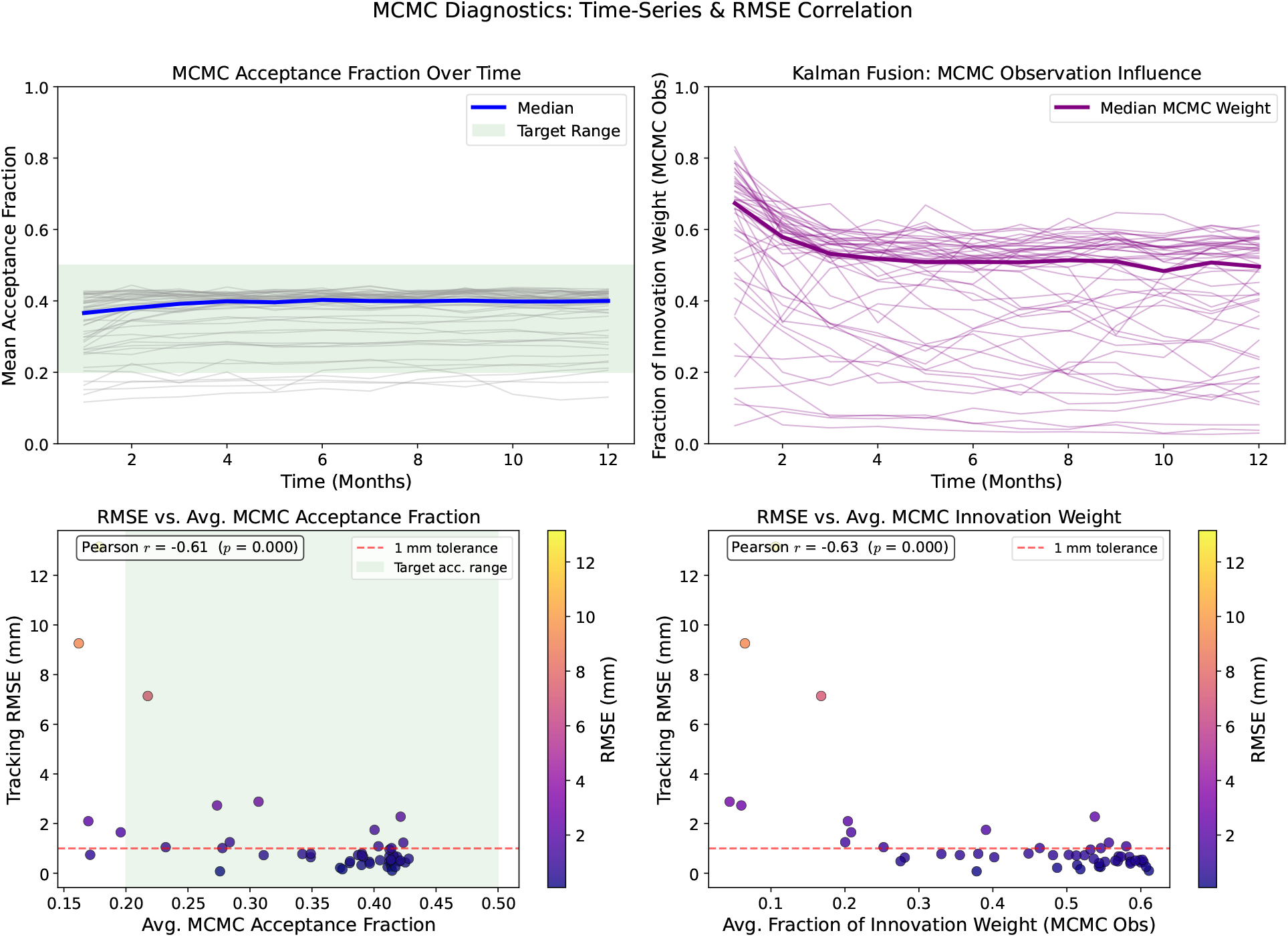
MCMC-Kalman tracking diagnostics across the patient cohort over a 12-month simulation period. (Top-left) Time-series of the mean MCMC acceptance fraction for each patient (gray traces) alongside the population median (blue). The green shaded band marks the target acceptance range of [0.2, 0.5] recommended for efficient ensemble MCMC sampling. (Top-right) Time-series of the fraction of Kalman filter innovation weight attributed to the MCMC-derived PPG likelihood observation (purple traces), with the population median in bold. Early in the tracking window, the MCMC observation dominates the update step (median ≈0.65), with the balance shifting toward the Kalman prediction as the state estimate matures and the process model becomes more informative. (Bottom-left) Scatter plot of per-patient tracking RMSE versus the time-averaged MCMC acceptance fraction, color-coded by RMSE magnitude. A statistically significant negative correlation (*r* = −0.61, *p <* 0.001) indicates that patients with higher MCMC target acceptance fraction achieved lower tracking errors. (Bottom-right) Scatter plot of tracking RMSE versus the time-averaged MCMC innovation weight. A similarly strong negative correlation (*r* = −0.63, *p <* 0.001) reveals that patients with higher MCMC innovation weight had lower tracking errors.

We reviewed the *θ*_circ_ estimation over time for 2 patients with high and low tracking RMSE values (0.51 mm vs. 2.10 mm) to examine whether parameter non-identifiability was coupled to diameter uncertainty or tracking error (Figure 8). In both cases, *θ*_circ_ was largely unidentified. However, in the patient with lower tracking RMSE, the relative standard deviation of *θ*_circ_ parameters generally reduced over time while in the patient with the larger RMSE, relative standard deviations of *θ*_circ_ parameters stayed constant over time or increased (Figure 8).

**Figure 8:**
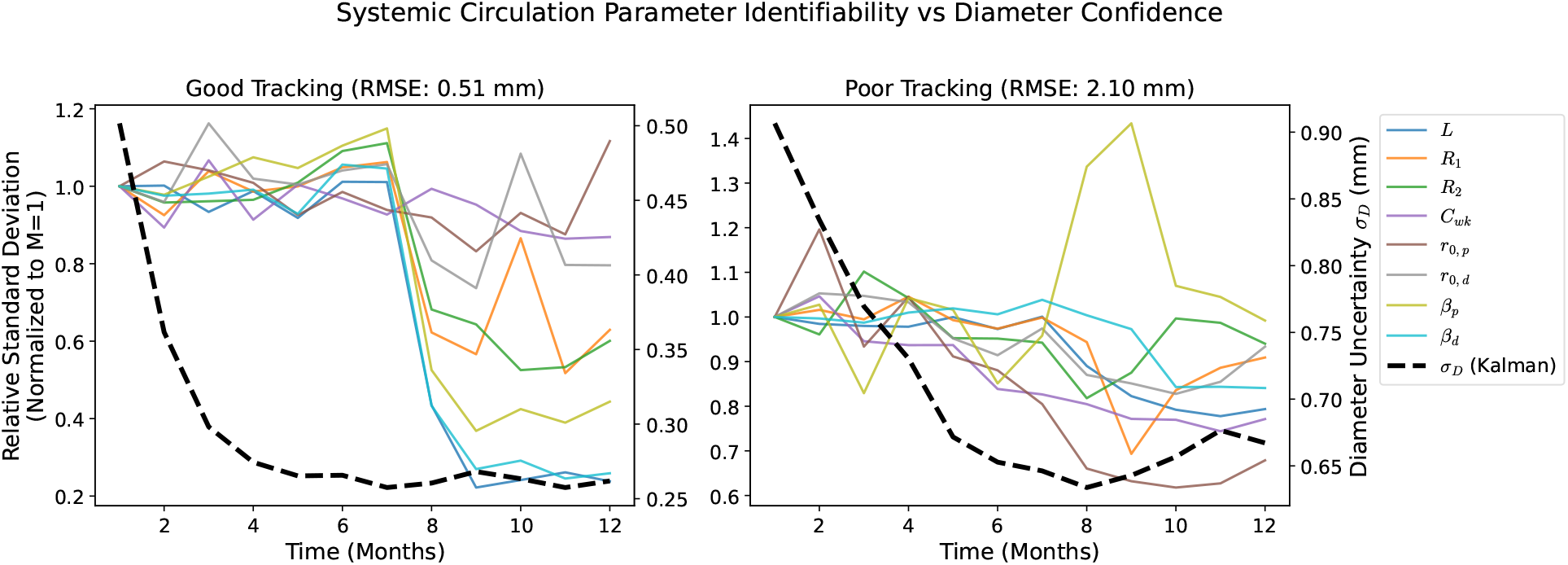
Systemic circulation parameter identifiability over time for representative patients with low and high tracking RMSE. Each panel shows the relative posterior standard deviation (left axis, normalized to the value at the first time step) of the eight systematic circulation parameters — vessel length *L*, peripheral resistances *R*_1_ and *R*_2_, Windkessel compliance *C*_*wk*_, proximal and distal reference radii *r*_0,*p*_ and *r*_0,*d*_, and wall stiffness coefficients *β*_*p*_ and *β*_*d*_ — inferred by the MCMC-Kalman tracker over a 12-month follow-up period. The Kalman diameter uncertainty *σ*_*D*_ (right axis, dashed black) is overlaid for reference.

## 4 Discussion

This work demonstrates the theoretical feasibility of tracking aortic aneurysm diameter using continuous PPG monitoring from wearable devices. Our physically-grounded, computational approach reveals both fundamental limitations and promising pathways for this novel application.

### 4.1 The Challenge of Single-Observation Estimation

Our sensitivity and identifiability analyses confirm an intuitive concern: detecting small geometric changes in the abdominal aorta from a peripheral measurement site is extraordinarily challenging. The condition numbers indicate that single PPG measurements cannot reliably distinguish diameter changes from physiological state variations in heart rate and blood pressure (Section 3.3 and Figure 1) or reliably identify systemic circulation parameters in the 1D model (Section 3.6). This finding aligns with prior work on pulse wave analysis, which has generally focused on proximal measurements or invasive sensors for aortic characterization (Nichols et al., 2011).

However, this limitation does not preclude clinical utility. The key insight is that wearable devices enable measurement paradigms impossible in clinical settings—specifically, the acquisition of thousands of observations per day under naturally varying physiological conditions. Moreover, these observations can be accumulated over weeks and months to refine posterior estimates of both the aneurysm diameter and the systemic parameters, as done in Section 3.7.

### 4.2 Exploiting Physiological Variation

The Cramér-Rao bound analysis demonstrates that aggregating observations across the natural range of heart rate and blood pressure effectively “separates” the diameter signal from confounding variables (Section 3.4). As HR and MAP vary throughout daily activities, the feature space is sampled along different directions, breaking the degeneracy that plagues single observations (Figure 2). This same approach holds true for estimating the systemic circulation parameters *θ*_circ_ with significant caveats due to the extreme condition number of the FIM and the strong coupling of uncertainty between *θ*_circ_ parameters (Section 3.6). This principle of aggregating measurements has been applied successfully in other wearable sensing domains, including PPG-based blood pressure estimation (Mukkamala et al., 2015).

The practical implication is that approximately 4 hours of cumulative monitoring (3,200 observations at 5 cardiac cycles each) can theoretically achieve sub-millimeter or millimeter-level diameter precision (Figure 3). For a wearable device worn continuously, this measurement window could be accumulated over hours to days, with the Bayesian fusion framework weighting observations by their uncertainty (Figures 4 and 6).

### 4.3 Robustness to Growth Dynamics

In Figures 4 and 6, the tracking simulations demonstrate that the Bayesian framework can detect and adapt to growth rate changes without explicit knowledge of acceleration events. This capability is clinically significant because sudden growth acceleration is precisely the scenario where continuous monitoring could provide the greatest benefit relative to intermittent imaging (Thompson et al., 2002).

The adaptive growth rate estimator (Section 2.5), while simple, proves sufficient to track the simulated growth patterns. More sophisticated approaches—such as change-point detection or growth model switching—could potentially improve response to abrupt transitions.

### 4.4 Parameter Identifiability and its Effect on Aneurysm Diameter Estimation

The FIM and CRB analysis on *θ*_circ_ parameters reveal a nuanced picture of parameter identifiability. While several parameters can hypothetically be estimated with reasonable accuracy from aggregating measurements (Figure 5), some may remain practically unidentifiable due to the extreme condition number and strong coupling of estimation uncertainty between parameters (Section 3.6). This finding is consistent with theoretical analyses of inverse problems in cardiovascular modeling (Banks et al., 2014; Nolte and Bertoglio, 2022).

Importantly, the sequential MCMC-Kalman based validation (Figure 6) with full posterior estimates of parameters achieves functional accuracy in aneurysm diameter tracking without needing to address the *θ*_circ_ parameter identifiability problem. The system exhibits robustness to certain parameter errors (Figure 8), suggesting that the combination of parameters most relevant to diameter sensitivity can be adequately characterized even when individual parameters are poorly constrained. Future work should investigate the connection between systemic circulation parameter identifiability and aneurysm diameter tracking accuracy more deeply.

Interestingly, Figure 7 suggests that specific MCMC-Kalman tracking diagnostics can be used to catch critical failures where aneurysm tracking errors are above threshold, without any ground-truth knowledge of aneurysm diameter or *θ*_circ_ parameters. There were some virtual patients with consistently low MCMC acceptance fraction and MCMC innovation weight fraction in Kalman fusion. These were generally the same cases with higher tracking RMSE values. An MCMC acceptance fraction and MCMC innovation weight < 0.25 seem to be indicative of tracking failure. This could be the result of over-confident initialization from the multi-start L-BFGS-B optimization. Such cases could be re-initialized with less confident priors or with a larger set of multi-starts to better sample the potential set of priors.

### 4.5 Clinical Translation Considerations

Several factors must be addressed for clinical translation. First, real PPG signals contain motion artifacts, varying skin contact, and environmental interference not present in our idealized simulations. Signal quality assessment and artifact rejection will be essential preprocessing steps. Second, our model assumes AAA location is known from initial imaging; detecting de novo aneurysms or aneurysms at atypical locations would require additional approaches. Third, all methods described assume access to continuous MAP measurements which can be feasibly derived from PPG features but is an active area of research (Mukkamala et al., 2015; Schrumpf et al., 2021).

The proposed workflow envisions PPG monitoring as a complement to, not replacement for, periodic imaging. Clinical imaging would provide ground-truth diameter measurements and re-calibration opportunities, while continuous PPG monitoring would interpolate between visits and flag unexpected changes for earlier follow-up. MCMC-Kalman diagnostic criteria could be used to flag patients for whom the tracker is likely failing without waiting until the next imaging visit. Corrective programmatic actions could be taken to re-initialize the tracking with better priors or make a recommendation for confirmatory imaging.

### 4.6 Limitations

This study has several limitations. First, the hemodynamic model, while physics-based, is a simplified 1D representation that omits three-dimensional flow features, vessel curvature, and branch effects. More sophisticated models (e.g., 3D-0D coupled simulations) would provide higher fidelity at increased computational cost (Reymond et al., 2009). Second, the neural surrogate, while accurate on the training distribution, may exhibit degraded performance for parameter combinations outside the training range. Third, the tracking simulations assume stationary noise characteristics; real wearable data quality varies with activity, posture, and device adherence. Fourth, we have not validated against clinical data; the simulated patients and growth patterns, while physiologically motivated, may not capture the full complexity of real AAA progression (Sherifova and Holzapfel, 2019).

## 5 Conclusion

We have demonstrated through computational analysis that continuous PPG monitoring could enable tracking of aortic aneurysm diameter between clinical imaging appointments. While single observations are fundamentally limited by poor identifiability, aggregating thousands of measurements under varying physiological conditions achieves millimeter-level precision according to Cramér-Rao bounds. Bayesian sequential estimation achieves sub-millimeter tracking when parameters are known and ∼1 mm tracking when they are not. Furthermore, runtime MCMC diagnostics–acceptance fraction and Kalman innovation weight–provide a means of detecting tracking failures without ground-truth imaging, enabling corrective action in a clinical workflow.

These results suggest that wearable PPG monitoring warrants further investigation as a complementary surveillance modality for aortic aneurysm patients. Clinical validation studies with expanded computational analysis correlating PPG-derived features with imaging-confirmed diameter changes are needed to translate these theoretical findings toward practical application.

## Data Availability

All data produced in the present study are available upon reasonable request to the authors

## A PPG Waveform Generation

Figure 9 in the main text demonstrates the physiological realism of simulated PPG waveforms. The model produces characteristic features observed in clinical PPG recordings: a rapid systolic upstroke reflecting the arrival of the pulse wave, a systolic peak followed by a brief notch (the dicrotic notch, representing aortic valve closure), and an exponential diastolic decay. As mean arterial pressure increases, the waveforms show expected changes including altered pulse pressure and modified dicrotic notch prominence.

**Figure 9:**
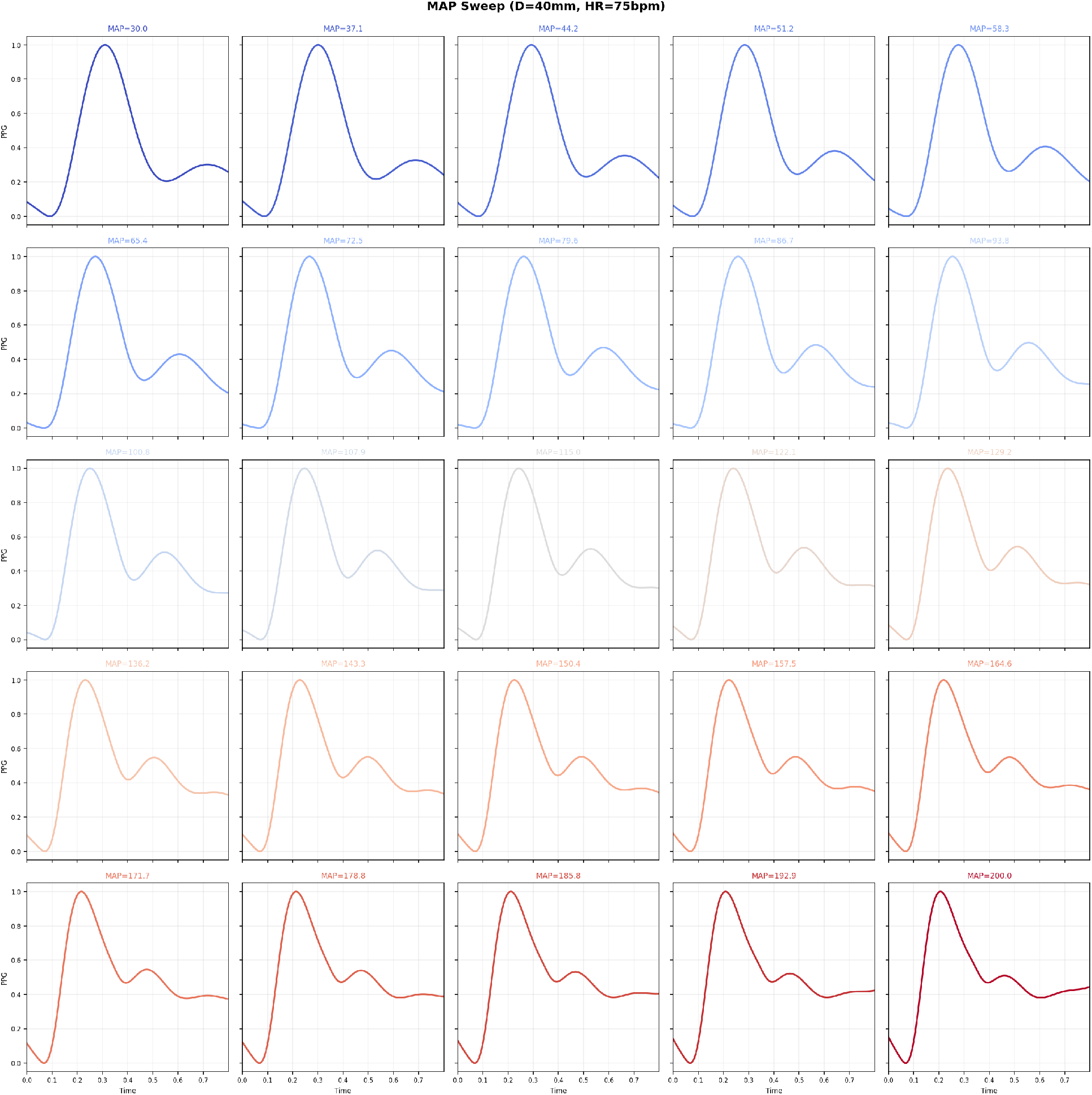
Simulated PPG waveforms at the pedal digital artery across a range of mean arterial pressures (30–200 mmHg) at fixed diameter (40 mm) and heart rate (75 bpm). Waveforms demonstrate physiologically realistic features including systolic peak, dicrotic notch, and diastolic decay. Color progression from blue (low MAP) to red (high MAP) illustrates systematic changes in waveform morphology with blood pressure.

## B Justification for MAP Measurement Uncertainty

We assumed *σ*_MAP_ = 10 mmHg for blood pressure estimation from PPG signals. This value is consistent with recent literature on cuff-less blood pressure monitoring. (Mukkamala et al., 2015) reported that PTT-based methods achieve approximately 8–12 mmHg error for systolic BP in controlled settings. (Schrumpf et al., 2021) found mean absolute errors of 7–15 mmHg for various PPG-based estimation approaches. We selected 10 mmHg as a representative value for current wearable technology, acknowledging that accuracy may improve with advances in calibration methods and sensor quality.

## C Neural Surrogate Performance

Table 2 provides detailed validation metrics for the neural surrogate model. Note: MAPE is not reported for AI due to values near zero causing numerical instability.

**Table 2:** Neural surrogate validation performance on held-out test set.

| Feature | RMSE | MAE | $R^2$ | MAPE (%) |
| --- | --- | --- | --- | --- |
| PTT (ms) | 5.22 | 2.57 | 0.76 | 5.0 |
| PWV<sub>eff</sub> (m/s) | 0.85 | 0.49 | 0.89 | 3.3 |
| AI | 0.057 | 0.024 | 0.89 | — |
| RI | 0.019 | 0.008 | 0.92 | 1.1 |
| SI (m/s) | 0.85 | 0.49 | 0.89 | 3.3 |
| $t_{\text{sys}}$ ratio | 0.018 | 0.008 | 0.95 | 2.4 |
| DN timing (ms) | 13.2 | 7.1 | 0.89 | 3.4 |

## D Fisher Information Matrix (FIM) structure for *θ*_circ_

Strong positive correlations (dark red) between *R*_1_–*R*_2_ and *L*–*β*proximal indicate that the PPG feature sensitivity directions of these parameter pairs are closely aligned (Figure 10, left), implying that their individual contributions to the observations are difficult to disentangle. The strong negative correlation between *β*_proximal_ and *β*_distal_ reflects a well-known opposing stiffness trade-off in Windkessel-type models.

**Figure 10:**
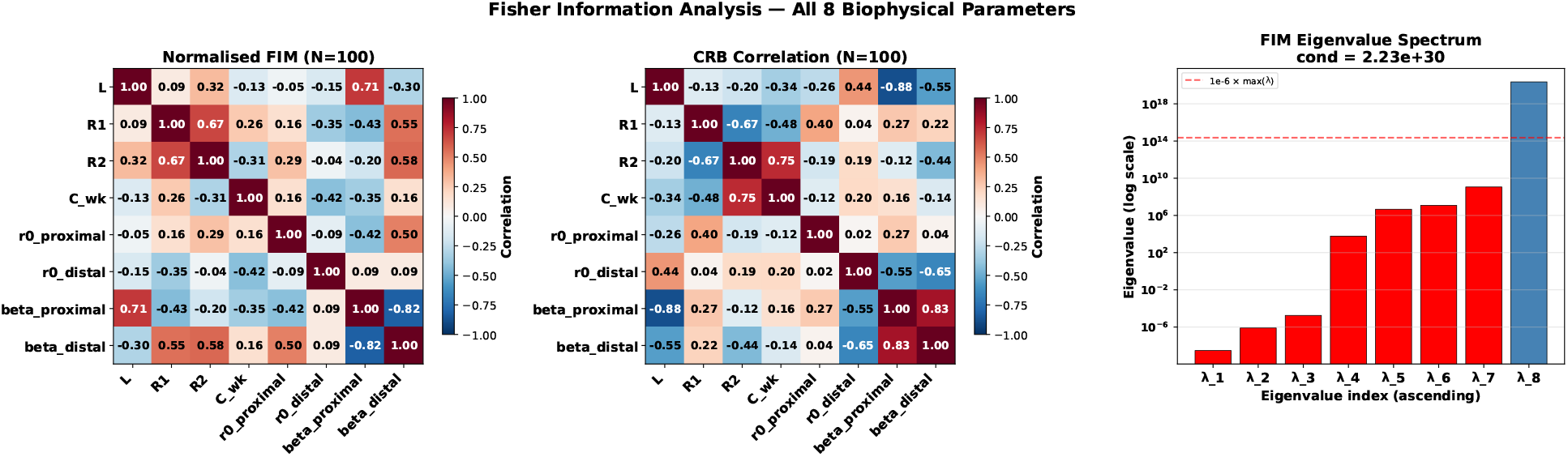
Fisher Information Matrix (FIM) structure for the eight systemic circulation parameters estimated from *N* = 100 PPG measurements. (Left) The normalized FIM, displayed as a correlation matrix, where entry (*j, k*) is 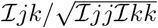. (Center) The Cramér-Rao Bound (CRB) correlation matrix, i.e., the off-diagonal structure of ℐ (***θ***)^−1^. (Right) The eigenvalue spectrum of the FIM (log scale), with a condition number of *κ* = 2.23 × 10^30^.

Large-magnitude off-diagonal entries—particularly between *L* and *β*_proximal_ (−0.88), *R*_1_ and *R*_2_ (−0.67), and *β*_proximal_ and *β*_distal_ (+0.83)—reveal strong estimation trade-offs (Figure 10, center); uncertainty in one parameter is systematically coupled to uncertainty in another, so errors cannot be independently reduced by increasing *N* alone.

The right-most panel of Figure 10 indicates severe near-singularity. The lowest seven eigenvalues (red) fall below the effective numerical rank threshold (10^−6^ ×*λ*_max_, dashed red line), confirming that the FIM is effectively rank-deficient at *N* = 100. This ill-conditioning means that certain linear combinations of parameters are essentially unobservable from the PPG feature space with a moderate measurement budget, motivating the use of larger *N* values while maintaining full posteriors during estimation along with imposing physiologically informed parameter constraints or process failure analysis during estimation.

